# Volumetric lung cancer screening reduces unnecessary low-dose computed tomography scans: results from a single-centre prospective trial on 4,119 subjects

**DOI:** 10.1101/2021.04.09.21255050

**Authors:** Gianluca Milanese, Federica Sabia, Roberta Eufrasia Ledda, Stefano Sestini, Alfonso Vittorio Marchianò, Nicola Sverzellati, Ugo Pastorino

## Abstract

**Purpose:** To compare low-dose computed tomography (LDCT) outcome and volume-doubling time (VDT) derived from measured volume (MV) and estimated volume (EV) of pulmonary nodules (PN) detected in a single-centre lung cancer screening trial.

**Materials and Methods:** MV, EV and VDT were obtained for prevalent pulmonary nodules detected at the baseline round of the bioMILD trial. LDCT outcome (based on bioMILD thresholds) and VDT categories were simulated on a PN- and a screenees-based analysis. Weighted Cohen’s kappa test was used to assess the agreement between diagnostic categories as per MV and EV.

**Results:** 1,583 screenees displayed 2,715 pulmonary nodules. On a PN-based analysis 40.1% PNs would have been included in different LDCT categories if measured by MV or EV. Agreement between MV and EV was moderate (κ = 0.49) and fair (κ = 0.37) for LDCT outcome and VDT categories, respectively.

On a screenees-based analysis, 46% pulmonary nodules would have been included in different LDCT categories if measured by MV or EV. Agreement between MV and EV was moderate (κ = 0.52) and fair (κ = 0.34) for LDCT outcome and VDT categories, respectively.

**Conclusions:** Within a simulated lung cancer screening based on recommendation by estimated volumetry, the number of LDCT performed for the evaluation of pulmonary nodules would be higher as compared to the prospective volumetric management.

## Introduction

Lung cancer screening (LCS) by Low-Dose Computed Tomography (LDCT) has been proved to reduce lung cancer (LC) mortality allowing early identification of pulmonary nodules (PN) with malignant behaviour. Beyond traditional risk factors, size and longitudinal growth drive management and diagnostic work-up of PNs detected on LCS (1-4). European LCS trials support volume as metric for assigning LDCT outcome, and the updated Lung CT screening Reporting and Data System (Lung-RADS 1.1) guidelines from the American College of Radiology (ACR) encompass volumetric thresholds for the assignment of diagnostic categories (5,6). Although the “true” volume of a PN may be of lesser relevance as compared to reproducibility of measurements - particularly during longitudinal evaluation - the management of prevalent PNs depends on size, as larger PNs require shorter time intervals between LDCT rounds (7). Measured volume (MV) of PNs obtained by dedicated software might have a positive impact on the sustainability of a LCS program by reducing the number of unnecessary LDCT rounds, as opposite to the geometrical conversion of diameters into the volume of a sphere (estimated volume, EV) - supported in the ACR Lung-RADS 1.1 guidelines, potentially leading to overestimated measurements (6,8).

We report the results of a simulation performed in a single-centre LCS trial, where MV is compared with EV for the assessment of both size and growth of solid PNs.

## Methods

This retrospective study was performed on the prospective data acquired from the baseline round of the bioMILD trial (clinicaltrials.gov ID: NCT02247453): a total of 4,119 subjects were prospectively enrolled at the “Istituto Nazionale dei Tumori” of Milan (Milan, Italy) between January 2013 and March 2016. LDCT scans were performed on a second-generation dual source CT scanner (Somatom Definition Flash; Siemens Medical Solutions, Forchheim, Germany), and MV was obtained by dedicated software (MM.Oncology, Syngo.via; Siemens Healthcare, Forchheim, Germany). Information on the bioMILD trial is detailed in *supplementary material*.

For the purposes of this study, we included solid PNs with a prospective software-measured maximum diameter of at least 3 mm. Diagnostic categories were assigned in keeping with the prospective thresholds of the bioMILD trial (*supplementary material*). First, each PN included in the electronic database was given a diagnostic category (PN-based analysis), and then the dominant PN was selected for each screenee (screenees-based analysis). EV was calculated from measurements derived from the semi-automated analysis.

Subsequently, we measured the volume doubling time (VDT) between baseline and first LDCT recall, on both PN- and screenees-based analyses. VDTs were stratified as follows: probably malignant PN (< 400 days), indeterminate PN (400 - 600 days), and probably benign PN (> 600 days).

### Statistical analysis

Data were expressed as percentages. Weighted Cohen’s kappa test was used to assess the agreement between diagnostic categories as per MV and EV. All statistical analyses were performed using Statistical Analysis System software, (version 9.4; SAS Institute, Cary, North Carolina, USA).

## Results

1,583 subjects (males: 65.5%) displayed 2,715 PNs. LDCT outcomes based on MV and EV are reported for the PN-based analysis in Table 1A and for the screenees-based analysis in Table 1B.

**Table 1A.** Low-dose Computed Tomography (LDCT) outcomes and volume doubling time (VDT) categories derived from measured volume (MV) and estimated volume (EV) for the PN-based analysis.

|  |  | PN-based analysis |  |  |
| --- | --- | --- | --- | --- |
|  |  | EV LDCT outcome |  |  |
|  |  | *Negative* | *Indeterminate* | *Positive* |
| MV LDCT outcome | *Negative* | 1308 | 735 | 160 |
|  | *Indeterminate* | 3 | 108 | 217 |
|  | *Positive* | 0 | 0 | 184 |
|  |  | PN-based analysis |  |  |
|  |  | EV VDT |  |  |
|  |  | *< 400* | *400 - 600* | *> 600* |
| MV VDT | *< 400* | 1021 | 29 | 254 |
|  | *400 - 600* | 29 | 12 | 11 |
|  | *> 600* | 377 | 73 | 505 |

**Table 1B.** Low-dose Computed Tomography (LDCT) outcomes and volume doubling time (VDT) categories derived from measured volume (MV) and estimated volume (EV) for the Screenees-based analysis.

|  |  | Screenees-based analysis |  |  |
| --- | --- | --- | --- | --- |
|  |  | EV LDCT outcome |  |  |
|  |  | *Negative* | *Indeterminate* | *Positive* |
| MV LDCT<br>outcome | *Negative* | 611 | 445 | 100 |
|  | *Indeterminate* | 1 | 83 | 182 |
|  | *Positive* | 0 | 0 | 161 |
|  |  | Screenees-based analysis |  |  |
|  |  | EV VDT |  |  |
|  |  | *> 600* | *400 - 600* | *> 600* |
| MV VDT | *< 400* | 579 | 15 | 170 |
|  | *400 - 600* | 17 | 5 | 5 |
|  | *> 600* | 225 | 40 | 291 |

### PN-based analysis

1,115 (40.1%) PNs would have been included in different LDCT categories if measured by MV or EV. VDT thresholds showed that 773 PNs out of 2,311 (33.4%) displayed different categories between MV-based and EV-based VDT.

Agreement between MV and EV was moderate (κ = 0.49) and fair (κ = 0.37) for LDCT outcome and VDT categories, respectively.

### Screenees-based analysis

728 (46%) PNs would have been included in different LDCT categories if measured by MV or EV (Figure 1A). VDT thresholds showed that 472 dominant PNs out of 1,347 (35.4%) displayed different categories between MV-based and EV-based VDT (Figure 1B).

**Figure 1A.**
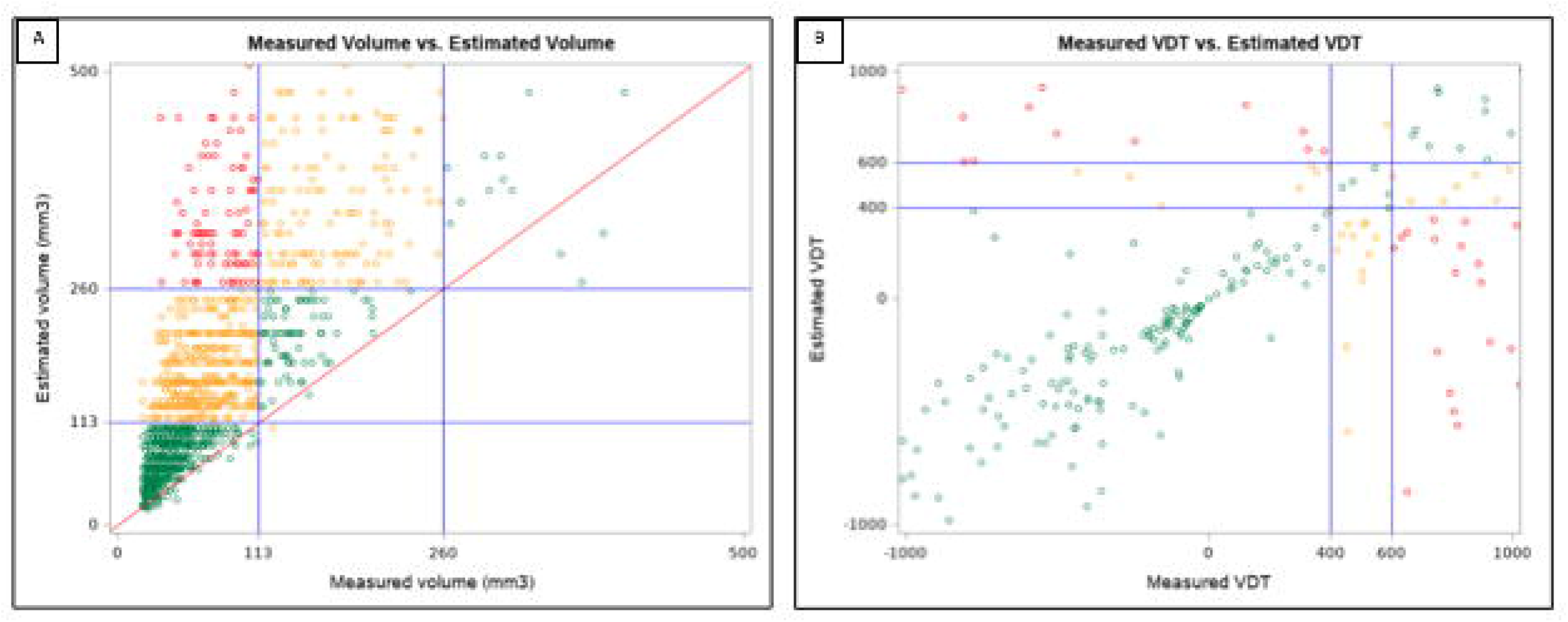
Box plot of measured volume and estimated volume for dominant pulmonary nodules, with the volumetric thresholds for negative and positive LDCT outcome highlighted. Only dominant pulmonary nodules with volumes ranging 3 and 500 mm^3^ are reported. (colour version online only) **Figure 1B.** Box plot of measured volume-VDT and estimated volume-VDT for dominant pulmonary nodules. Only dominant pulmonary nodules with VDT within −1,000 and 1,000 are reported. (colour version online only)

Agreement between MV and EV was moderate (κ = 0.52) and fair (κ = 0.34) for LDCT outcome and VDT categories, respectively.

## Discussion

We compared measured volume and estimated volume of solid PNs detected at the baseline round of a single-centre lung cancer screening trial: discrepancies between MV-based and EV-based evaluation resulted in different post-test diagnostic categories and different growth assessment, with a higher number of LDCT recalls and invasive testing from the EV-based analysis.

Volumetry of PNs has been adopted in a number of European LCS trials, and its measurement has been recommended also for the management of PNs in the daily practice by the Fleischner Society and the British Thoracic Society (9-11). Indeed, MV has been recognized to be more reproducible as compared to diameter measurement, which suffers from limited intra- and inter-reader agreement (12). However, beyond the definition of the most accurate method, one major question is whether one method better predicts prognosis and outcome than another (8). Indeterminate PNs are expected in about 10-20% of screenees: in our analysis, the percentage was 16.9% for the MV-based analysis, and 33.4% for the EV-based one. Such higher number of EV-based indeterminate PNs should be investigated from a clinical perspective to verify whether it includes malignant lesions or if it is due to false-positive lesions. However, such analysis, albeit of paramount relevance (8), goes beyond the scope of this study. We look forward to the results of the bioMILD trial that may answer the question on personalized screening intervals based on a combination of LDCT outcome and individual risk profiling by plasma miRNA analysis.

On a PN-based analysis, the 58.9% of PNs would have been included within the same LDCT outcome: EV would have caused an increased number of LDCT recalls, particularly for indeterminate PNs. Agreement between LDCT outcome and between VDT categories for both MV and EV was moderate and fair, respectively, with only a minor increase in the coefficient of agreement for screenees-based analysis as compared with the PN-based analysis. Risk stratification by MV might improve the affordability of LCS for national health systems, as it could grant a significant reduction of LDCT recalls throughout the LCS duration. The recently updated LungRADS categories added volumetric thresholds to diameter thresholds, and these outcome categories – assigned from semi-automated volumetric measurements of prevalent PN - could predict LC risk for screenees enrolled in the MILD trial that fulfilled NLST selection criteria, supporting a two-years interval for negative LDCT outcome, and by reducing the number of LDCT to performed, such approach might increase sustainability of LCS (13). Notably, in our screenees-based analysis, the 99.9% of the differently classified PNs by EV would have requested a shorter follow-up as compared with the MV approach, and only three PNs would have been recalled following a 3-year (EV) rather than a 12-months interval (MV). Same results would have been derived after evaluation of EV-based VDT, a metric that is of paramount relevance for management of PNs, either diameter-based or volumetry-based. As a variation of 20% in average diameter equates with a variation in volume of about 100%, slight differences could result in growth rate, requiring invasive testing for confirmation of the biological behaviour of PNs (9). This would have led to higher costs for the radiology departments and, from a screenees perspective, to higher radiation exposure, higher risks from invasive manoeuvres and anxiety for a greater proportion of subjects, which may cause psychological burden limiting engagement and adherence to LCS (5).

We do recognize that PNs are not necessarily spherical and that EV - converting the maximum diameter into a sphere - will intrinsically cause an overestimation of the volume of PNs. Although we did not include the mean diameter, and only tested the maximum diameter obtained by a semi-automated software to limit the low reproducibility of manual measurements, our results are in keeping with those from the NELSON trial (14,15). In daily clinical practice, however, only a minority (8%) of radiologists reported to use volumetric software (16). We aimed to test the impact of the geometrical conversion of a diameter into the volume of a sphere on the management of PNs. Such conversion could represent an alternative to MV, particularly in those setting where such tools are not available, being follow-up timing strictly dependent on lesion measurement. Our EV-approach could be considered an extension of the diameter-based approach of the NLST (e.g., LDCT positive outcome in case of any diameter greater than a specified threshold) (1). Furthermore, the software defines the maximum diameter with a three-dimensional approach, which may outperform the measurements obtained on axial images, which is further more susceptible to a reader-based decision on the slice were the maximum diameter is displayed for its manual measurement. Potential implementation of machine-learning approaches (e.g., removal of vascular structures for accurate outlining boundaries of PNs; radiomics analysis suggesting the risk of malignancy of PNs) might modify the management of PNs detected on LCS in the future, reducing the impact of readers intervention toward the measurement process (e.g., manual correction of inaccurate semi-automated segmentations) (17).

One strength of our study is the single centre setting, with LDCT acquired with one single scanning protocol on a dedicated CT scanner; on the other hand, such setting might limit generalizability of the results. Hence, we foster future studies comparing EV and MV in a multicentric and multivendor design. Moreover, image interpretation was performed with one software throughout the trial, allowing reduction of inter-scanner and inter-software variability. On the other hand, we could not test the impact of different acquisition parameters, reconstruction algorithms and software, which might influence the results of the semi-automated volumetric analysis. Our study presents some limitations. Our analyses are not stratified by nodule morphology and we excluded sub-solid PNs, as the version of the software available at the time of the prospective reading did not allow their semi-automated volumetry analysis. We foster future studies testing accuracy of newer versions of the software. As previously discussed, the results of EV might be overestimated as we considered PN as spherical structures and did not evaluate average diameters. Results of semi-automated segmentations were prospectively collected, and information on the number of manual corrections of the PNs’ contours that were needed were not evaluated in the present study.

In conclusion, within a simulated LCS based on recommendation by estimated volumetry, the number of LDCT performed for the evaluation of PNs would be higher as compared to the prospective volumetric management.

## Supporting information

COI Disclosure AM

COI Disclosure FS

COI Disclosure GM

COI Disclosure NS

COI Disclosure REL

COI Disclosure SS

COI Disclosure UP

## Data Availability

Data will not be made available.

