## Supplemental Materials for "Volumetric lung cancer screening reduces unnecessary low-dose computed tomography scans: results from a single-centre prospective trial on 4,119 subjects"

### Supplementary materials

#### Study population

This retrospective study was performed on the data from the baseline round of the bioMILD trial, a large prospective study testing the combination of plasma miRNA and LDCT to improve the efficacy of LCS by individual risk profiling and personalised screening intervals (clinicaltrials.gov ID: NCT02247453).

#### Scanning protocols

The whole chest volume was scanned during one deep inspiratory breath-hold, with the following scanning parameters: tube voltage 120 kVp, tube current 30 mAs, collimation 0.625 mm, pitch 1.2, rotation time 0.5 seconds. Images were reconstructed with the following parameters: thickness 1 mm; increment 0.7 mm; medium-sharp kernel (B50f); lung window setting (window width 1600 HU, window level −600 HU).

#### LDCT outcome

Diagnostic categories were assigned in keeping with the prospective thresholds of the bioMILD trial:

1. Negative outcome: PN <113 mm^3^
2. Indeterminate outcome: PN 113 - 260 mm^3^
3. Positive outcome: PN >260 mm^3^

#### Statistical analysis

Diameter-based nodule volume was calculated using the maximal diameter by assuming a spherical nodule shape (formula: $V=\frac{1}{6}\times\pi\times D^{3}$, with V = volume and D = maximal diameter).

VDT was calculated using the following formula: $VDT= \frac{[\ln2 \times\Delta T]}{[\ln(\frac{Volume T2}{Volume T1})]}$, where $\Delta T$ is the time (in days) between the two scans.
